# Real-world experience with molnupiravir during the period of SARS-CoV-2 Omicron variant dominance

**DOI:** 10.1101/2022.07.05.22277227

**Authors:** Robert Flisiak, Dorota Zarębska-Michaluk, Magdalena Rogalska, Justyna Anna Kryńska, Justyna Kowalska, Ewa Dutkiewicz, Krystyna Dobrowolska, Jerzy Jaroszewicz, Anna Moniuszko-Malinowska, Marta Rorat, Regina Podlasin, Olga Tronina, Piotr Rzymski

**Author notes:** **The corresponding author:** prof. dr hab. Robert Flisiak, 15-540 Białystok, ul. Żurawia 14, Poland; e-miał. equal contribution (Robert Flisiak, Dorota Zarębska-Michaluk).

## Abstract

**Background:** The real-world effectiveness of molnupiravir (MOL) during the dominance of Omicron SARS-CoV-2 lineage is urgently needed since the available data relates to the period of circulation of other viral variants. Therefore, this study assessed the efficacy of MOL in patients hospitalized for COVID-19 in a real-world clinical practice during the wave of Omicron infections.

**Methods:** Among 11822 patients hospitalized after 1 March 2020 and included in the SARSTer national database, 590 were treated between 1 January and 31 April 2022, a period of dominance of the Omicron SARS-CoV-2 variant. MOL was administered to 203 patients, whereas 387 did not receive any antiviral regimen. Both groups were similar in terms of sex, BMI and age allowing for direct comparisons

**Results:** Patients who did not receive antiviral therapy significantly more often required the use of Dexamethasone and Baricitinib. Treatment with MOL resulted in a statistically significant reduction in mortality during the 28-day follow-up (9.9 vs. 16.3%), which was particularly evident in the population of patients over 80 years of age treated in the first 5 days of the disease (14.6 vs. 35.2%). MOL therapy did not affect the frequency of the need for mechanical ventilation, but patients treated with MOL required oxygen supplementation less frequently than those without antivirals (31.7 vs. 49.2%). The time of hospitalization did not differ between groups.

**Conclusions:** The use of molnupiravir in patients hospitalized for COVID-19 during the dominance of Omicron variant reduced mortality. This effect is particularly evident in patients over 80 years of age.

## Introduction

The first infections of the novel coronavirus 2019 (2019-nCov), later named severe acute respiratory syndrome coronavirus 2 (SARS-CoV-2), a causative agent of the coronavirus disease 2019 (COVID-19), were documented in China in December 2019. The rapid spread of the disease and an increasing number of infections prompted World Health Organization (WHO) to declare COVID-19 a pandemic as early as March 2020. The clinical spectrum of respiratory infections ranges from mild to critical, with several percent of patients dying from respiratory failure, shock, and multiple organ failure. Due to such consequences, the number of cases quickly made the disease one of the major sources of morbidity and mortality worldwide. Virus genetic variability posed an additional problem, as successive variants differed not only in their infectivity and pathogenicity but also in the effectiveness of the drugs used against them. This became most apparent with the currently dominant Omicron variant [1,2].

Since the beginning of the pandemic, an urgent need for effective forms of therapy has arisen to reduce this global health burden. Due to the lack of time needed to create a new drug, the researchers’ attention has focused on using existing drugs for a new indication. Thus, MOL, an antiviral drug originally developed to treat patients with Venezuelan equine encephalitis virus infection and later, in the pre-pandemic period, entered preclinical studies with influenza, has come into the spectrum of attention [3–5]. MOL, a polymerase inhibitor prodrug that acts as a synthetic nucleoside, is administered orally, an advantage over the previously available intravenous, antiviral drug, remdesivir, used to treat COVID-19 and facilitates therapy in the out-of-hospital setting [6]. It was the phase 2/3 MOVe-OUT clinical trial in non-hospitalized patients with SARS-CoV-2 infection that was the basis for the Food and Drug Administration (FDA) December 2021 approval of MOL for emergency use in the treatment of mild to moderate COVID-19 virus infection in adults at high risk of progression to severe disease [7]. No clinical benefit was seen in the MOVe-IN clinical trial evaluating MOL in hospitalized patients [8]. This study, however, was conducted in the period of dominance of the earlier SARS-CoV-2 variants and does not necessarily reflect the current situation related to the dominance of the Omicron variant.

The purpose of this analysis was to assess the efficacy of MOL in patients, particularly the elderly, hospitalized for COVID-19 in a real-world clinical practice during the period of the Omicron variant dominance.

## Patients and Methods

Patients included in the analysis originated from the SARSTer national database, which included 11822 patients treated between 1 March 2020 and 31 April 2022 in 30 Polish centers. This ongoing project, supported by the Polish Association of Epidemiologists and Infectiologists (PTEiLChZ), is a national real-world experience study assessing treatment in patients with COVID-19. The study was conducted according to the guidelines of the Declaration of Helsinki, and approved by the Ethics Committee of the Medical University of Białystok (29 October 2020, number APK.002.303.2020). All the patients were diagnosed with COVID-19 based on positive results of the real-time reverse transcriptase-polymerase chain reaction (RT-PCR) or using an antigen test from the nasopharyngeal swab specimen. The decision about the treatment regimen was taken entirely by the treating physician concerning current knowledge and recommendations of the PTEiLChZ [9,10]. The present study included 590 adult patients hospitalized between 1 January and 31 April 2022, which is considered the period of dominance of the Omicron SARS-CoV-2 variant in Poland [11]. Molnupiravir was administered orally twice daily at 800 mg for 5 days to 203 patients according to PTEiLChZ recommendations, whereas 387 did not receive any antiviral regimen [9,10]. Data were entered retrospectively and submitted online by a web-based platform operated by Tiba sp. z o.o. Baseline data included age, gender, body mass index (BMI), coexisting conditions, use of other COVID-19 medications, clinical and laboratory measures including C-reactive protein (CRP), procalcitonin, white blood cells (WBC), platelets, interleukin 6 (IL-6), and d-dimers. Treatment effectiveness end-points were 28 days mortality rate and the need for mechanical ventilation. Moreover, clinical improvement was analyzed with ordinal scale categories at consecutive time points on day 7, 14, 21, or 28 depending on baseline oxygen saturation (SpO2), patient’s age and molnupiravir administration within 5 days of symptom onset. This scale is based on WHO recommendations modified to fit the specificity of the national health care system as applied previously [12,13]. The ordinal scale was scored as follows: 1) unhospitalized, no activity restrictions; 2) unhospitalized, with limited activity; 3) hospitalized, does not require oxygen supplementation or medical care; 4) hospitalized, requiring no oxygen supplementation, but requiring medical care; 5) hospitalized, requiring normal oxygen supplementation; 6) hospitalized, on non-invasive ventilation with high-flow oxygen equipment; 7) hospitalized, on invasive mechanical ventilation or extracorporeal membrane oxygenation (ECMO); 8) death.

## Statistical analyses

The results were expressed as mean ± standard deviation (SD) or n (%). All statistical calculations were performed with Statistica v. 13.3 (StatSoft, USA). Since the data distribution did not meet the Gaussian assumption, non-parametric methods were applied to assess the difference between MOL and NO AVT groups. Differences in frequencies in parameters given as nominal categorical variables were evaluated with Pearson’s χ^2^ test. A p-value < 0.05 was considered statistically significant.

## Results

As shown in Table 1, the study groups were similar in terms of sex, BMI and age, with as many as 73% of patients treated with MOL and 71% of the untreated group over 60 years of age. Except for platelet count, there were no significant differences between the groups of patients in the baseline SpO2 and values of major laboratory indicators used in monitoring the course of COVID-19 (Table 1). Patients who did not receive antiviral therapy significantly more often required the use of immunomodulating drugs during 28 days follow-up period, especially dexamethasone and baricitinib (Table 1).

**Table 1.**
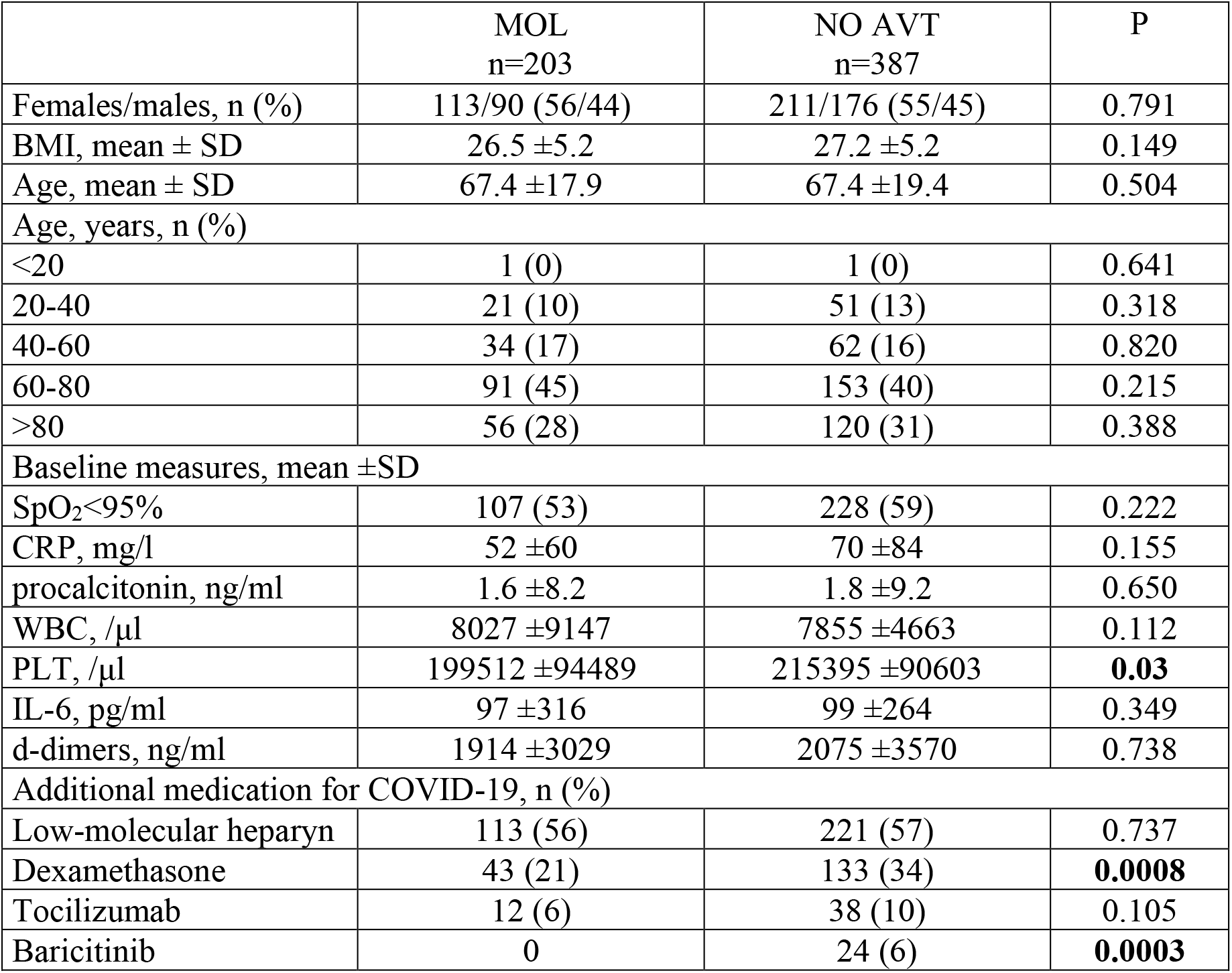
Clinical characteristics of all adult patients hospitalized between 1-01-2022 and 31-04-2022 receiving molnupiravir (MOL) or no antiviral therapy (NO AVT).

A group of patients who received MOL within the first 5 days of the disease was characterized from the 7th day of disease by the tendency for a lower percentage of deaths (5.1 vs. 9.2%, p=0.05), the need for oxygen therapy (42.4 vs. 50.4, p=0.06) and a significantly higher percentage of hospital discharge (12.1 vs. 5.7%, p=0.005) (Figure 1). The differences in the death rates between groups (5.7 vs. 10.3%, p=0.01) at day 7 increased in patients over 60 years of age (Figure 2), and especially in those over 80 (7.3 vs. 16.5%, p=0.03), (Figure 3). As shown in Table 2, treatment with MOL resulted in a statistically significant reduction in mortality during the 28-day follow-up. This was particularly evident in the population of patients over 80 years of age who took the drug in the first 5 days of the disease. However, antiviral therapy did not affect the frequency of the need for mechanical ventilation (Table 2).

**Table 2.** Twenty-eight days mortality and the need for mechanical ventilation in patients treated with molnupiravir (MOL) or without any antiviral treatment (NO AVT) analyzed in all patients, only those with oxygen saturation ≤95%, including administration of molnupiravir within 5 days of symptom onset as well as aged over 60 or 80 years.

| Patients subpopulations | Mortality |  |  | Mechanical ventilation |  |  |
| --- | --- | --- | --- | --- | --- | --- |
|  | MOL<br>n/N (%) | NO AVT<br>n/N (%) | P | MOL<br>n/N (%) | NO AVT<br>n/N (%) | p |
| All patients | 20/203<br>(9.9) | 63/387<br>(16.3) | **0.03** | 7/203<br>(3.5) | 14/387<br>(3.6) | 0.916 |
| SpO<sub>2</sub>$\leq 95\%$ | 17/107<br>(15.9) | 52/228<br>(22.8) | 0.114 | 5/107<br>(4.7) | 11/228<br>(4.8) | 0.952 |
| SpO<sub>2</sub>$\leq 95\%$,<br>0-5 days, | 14/99<br>(14.1) | 52/228<br>(22.8) | 0.073 | 3/99<br>(3.0) | 11/228<br>(4.8) | 0.461 |
| SpO<sub>2</sub>$\leq 95\%$,<br>0-5 days,<br>>60 years. | 14/88<br>(15.9) | 49/195<br>(25.1) | 0.08 | 3/88<br>(3.4) | 11/195<br>(5.6) | 0.423 |
| SpO<sub>2</sub>$\leq 95\%$,<br>0-5 days,<br>>80 years. | 6/41<br>(14.6) | 32/91<br>(35.2) | **0.016** | 0/41<br>(0) | 3/91<br>(3.3) | 0.239 |

**Figure 1.**
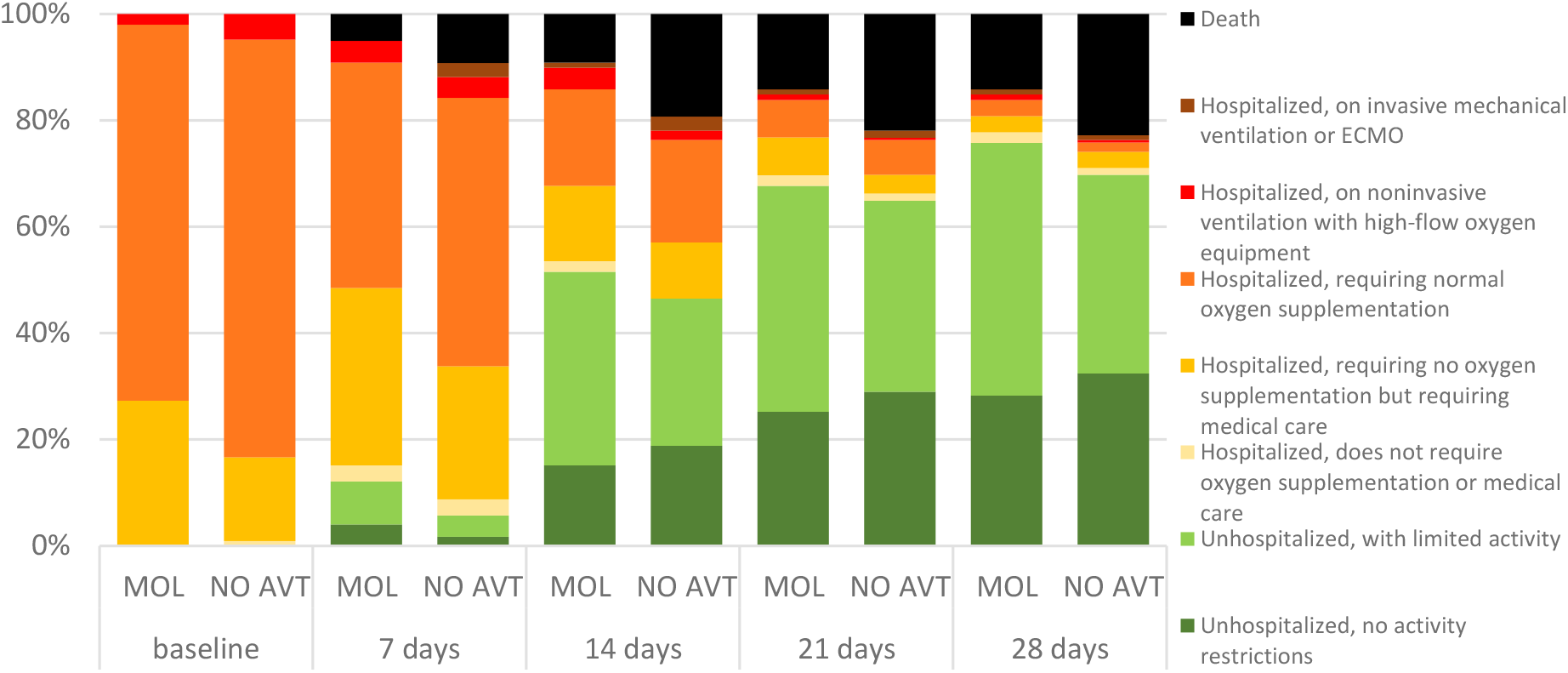
Ordinal scale categories at consecutive time points in all patients with SpO2≤95% treated with molnupiravir (MOL) within 5 days of symptom onset or no antiviral treatment (NO AVT).

**Figure 2.**
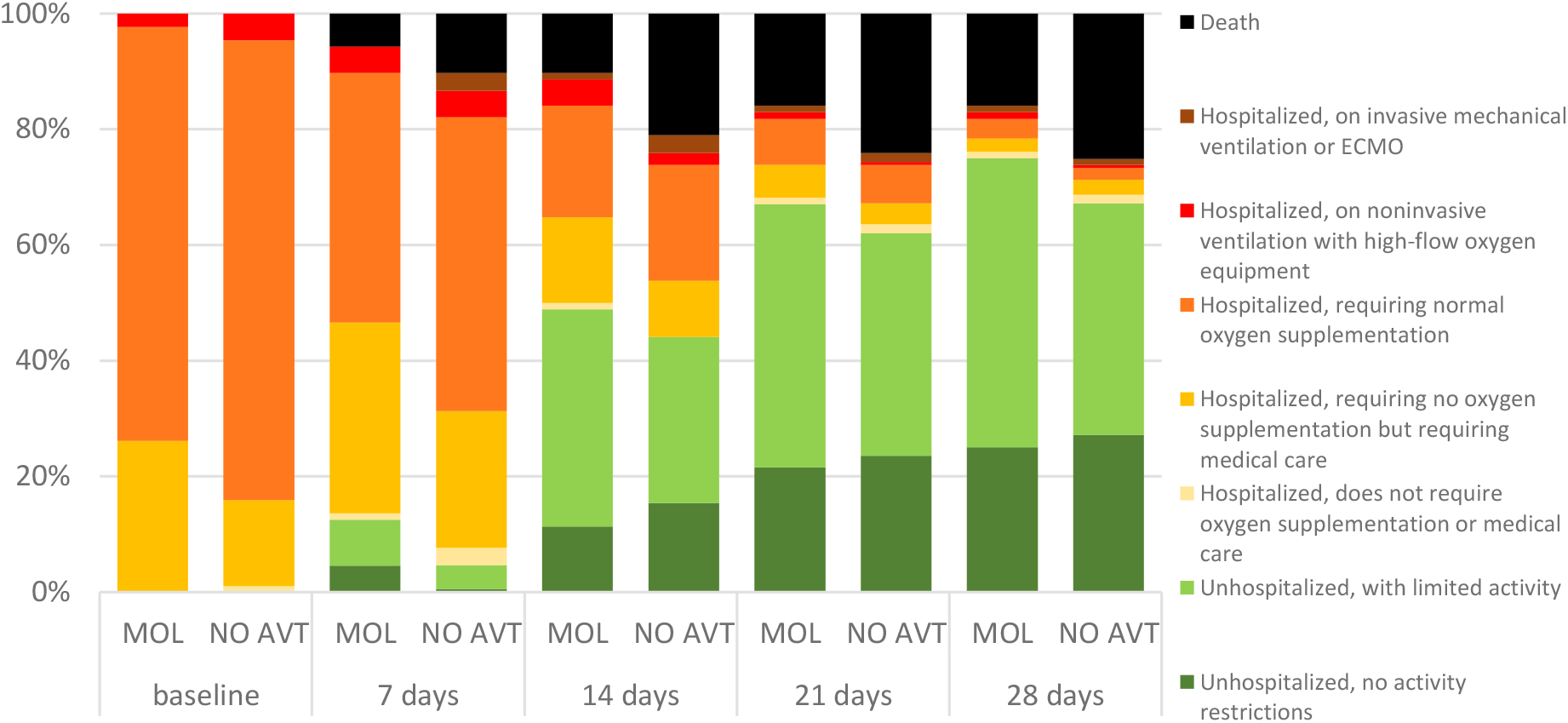
Ordinal scale categories at consecutive time points in patients over 60 years of age with SpO2≤95% treated with molnupiravir (MOL) within 5 days of symptom onset or no antiviral treatment (NO AVT).

**Figure 3.**
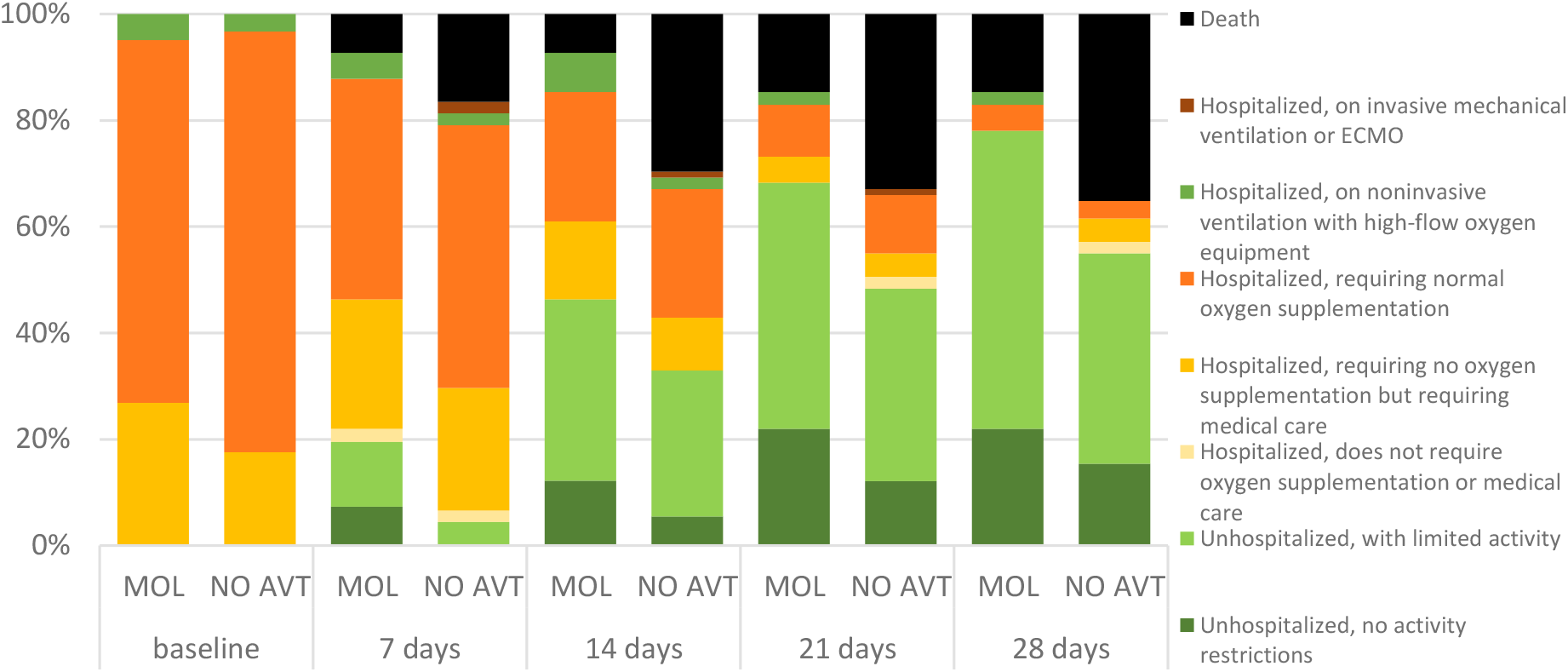
Ordinal scale categories at consecutive time points in patients over 80 years of age with SpO2≤95% treated with molnupiravir (MOL) within 5 days of symptom onset or no antiviral treatment (NO AVT).

The time of hospitalization did not differ between patients treated with MOL and without any antiviral treatment (mean±SD 11.6±7.9 vs. 11.5±9.3 days; p=0.965), and this lack of difference was also observed in the subset of patients aged >60 years (14.1±9.1 vs. 13.9±9.3 days; p=0.679) and 80 years (14.2±9.6 vs. 14.01±11.4 days; p=0.855). All patients treated with MOL during the whole 28-days follow-up period required oxygen supplementation less frequently than those not treated with any antivirals (31.7 vs. 49.2%, p=0.00005). This difference was also evident in the subset of patients aged > 60 years (56.3 vs. 73.8%, p=0.003) but not in the subset of patients >80 years (62.5 vs 69.3%, p=0.449).

## Discussion

Although the MOVe-IN clinical trial evaluating the use of MOL in a population of hospitalized patients was discontinued due to a lack of expected benefits, data from both the MOVe-OUT and MOVe-IN studies demonstrated that MOL appears to inhibit replication of the virus and is the most effective when therapy is started early in the disease course in patients with mild-to-moderate COVID-19 [7,8]. The consequence of discontinuing the clinical trial in hospitalized patients was that the FDA did not issue an emergency use authorization of MOL in this population, and there was a paucity of reports of treatment efficacy in such patients in the real-world population, making our analysis unique. To date, the only study from routine clinical practice on the use of MOL in hospitalized patients has been published as a preprint and presents the experience of a center in Hong Kong, confirming the efficacy of oral antivirals, including MOL used in 2116 patients, in reducing mortality and risk of disease progression [14].

Therefore, the current multicenter study aimed to compare hospitalized COVID-19 patients treated with MOL with those who did not receive antiviral therapy to determine the potential clinical benefit, filling this knowledge gap. It is important to note that both our analysis and the study performed by Carlos et al. included patients hospitalized during the pandemic wave of the Omicron variant [14], whereas recruitment to the MOVe-IN study occurred during a period of the predominance of previous SARS-CoV-2 variants of concern, Alpha and Delta [7].

Despite the altered sensitivity of the Omicron mutation to the vaccines and some COVID-19 therapeutics, data available from in vitro studies indicate that the antiviral drugs, remdesivir, nirmatrelvir/ritonavir, and MOL remain active against this variant of concern [2]. The study population consisted of patients requiring hospitalization due to COVID-19 and burdened with risk factors of severe disease course related to comorbidities and age; 73% were patients over 60 years of age, whereas in the MOVe-IN study, such patients accounted for only 42% [7]. Also noteworthy is the significantly higher proportion of women in our analyzed group compared to the MOVe-IN study (56% vs. 43%), which may have influenced the result.

The present analysis demonstrated that the use of MOL was associated with a significant reduction in all-cause mortality compared to using no antiviral drugs, 9.9% vs. 16.3% (p=0.03). Thus, our data are consistent with the results obtained by Carlos et al., who reported a mortality rate in the MOL-treated group of 8.9% versus 15.9% in the control group (p<0.001) [14]. In our study, the difference was most pronounced for the subset of patients aged over 80 years with baseline oxygen saturation ≤95% who received MOL within the first 5 days from the onset of symptoms, 14.6% vs. 35.2% (p=0.016). Due to the fact that the baseline indices of the disease activity did not differ significantly in the two groups, the higher frequency of immunomodulating drugs among patients not treated with antivirals should be considered as reflecting the progression of the disease later in the course of the disease.

We found no statistically significant differences in terms of the need for mechanical ventilation, in contrast to a report from Hong Kong, which showed a significantly lower risk of this event in patients treated with MOL compared to the group not treated with antivirals, 0.4% vs. 1.4% (p<0.001) [14]. These discrepancies in the results may have been influenced by the difference in the size of the cohorts analyzed, 203 vs. 2116 patients. It is noteworthy, however, that in our study none of the 41 patients older than 80 years with baseline oxygen saturation ≤95% who began MOL therapy within the first 5 days required mechanical ventilation, contrary to 3 out of 91 patients in the group not receiving antiviral therapy. The finding of a statistical association between the timing of MOL inclusion and its efficacy in the MOVe-OUT and MOVe-IN trials became the basis for recommending its use up to five days after the onset of disease symptoms [7–9].

In the population of patients over 80 years of age with baseline saturation ≤95% who received therapy within the first five days, the trend of more rapid clinical improvement as assessed by the WHO ordinal scale was also most evident, although this association was demonstrated in the entire group of patients receiving MOL compared with those not receiving antiviral treatment.

We are aware that although our study provides a valuable addition to the knowledge of MOL use in patients with COVID-19, it has some limitations related to its real-world and multicenter retrospective observational nature with possible bias and entry errors. However, its real-world nature is also the strength of the study, as the analyzed population is heterogeneous, covering different parts of the country, which allows the results to be generalized.

In conclusion, we have documented that the use of MOL during the first five days in patients hospitalized for COVID-19 and burdened with risk factors for severe disease significantly reduces mortality and contributes to clinical improvement. This relationship is particularly evident in patients over 80 years of age and baseline oxygen saturation ≤95%.

## Data Availability

All data produced in the present study are available upon reasonable request to the authors

## Abbreviations

COVID-19: coronavirus disease 2019
CRP: C-reactive protein
ECMO: extracorporeal membrane oxygenation
FDA: Food and Drug Administration
NO AVT: no antiviral therapy
IL-6: interleukin 6
MOL: molnupiravir
PTEiLChZ: Polish Association of Epidemiologists and Infectiologists
RT-PCR: real-time reverse transcriptase–polymerase chain reaction
SARS-CoV-2: severe acute respiratory syndrome coronavirus 2
SpO2: oxygen saturation
WBC: white blood cells
WHO: World Health Organization,

## Conflict of interest statement in relation to the work

Robert Flisiak and Jerzy Jaroszewicz reports grants and personal fees from MSD, Gilead, and Roche; Dorota Zarębska-Michaluk reports personal fees from MSD and Gilead; Justyna Kowalska reports grants and personal fees from MSD, Gilead, Janssen Cilag, GSK/ViiV and Roche; Anna Moniuszko-Malinowska reports personal fee from DiaSorin; Regina Podlasin reports personal fees from Gilead; Magdalena Rogalska, Justyna Anna Kryńska, Ewa Dutkiewicz, Krystyna Dobrowolska, Marta Rorat, Olga Tronina, Piotr Rzymski report no financial interests in relation to the work described

## Data Availability Statement

the datasets generated during and/or analysed during the current study are available from the corresponding author on reasonable request.

## Author contributions (mandatory) for all authors

Robert Flisiak – concept and design of the study, data analysis, wrote part of the article, drawn figures, performed the final submission; Dorota Zarębska-Michaluk - data collection and analysis, wrote part of the article, prepared tables, final correction of the manuscript; Piotr Rzymski – data analysis, statistics, wrote part of the article; Magdalena Rogalska, Justyna Anna Kryńska, Justyna Kowalska, Ewa Dutkiewicz, Krystyna Dobrowolska, Jerzy Jaroszewicz, Anna Moniuszko-Malinowska, Marta Rorat, Regina Podlasin, Olga Tronina – data collection and analysis. All authors have read and approved the final version of the manuscript.

## References

[1] Mader A-L, Tydykov L, Glück V, Bertok M, Weidlich T, Gottwald C, et al. Omicron’s binding to sotrovimab, casirivimab, imdevimab, CR3022, and sera from previously infected or vaccinated individuals. IScience 2022;25:104076.

[2] Vangeel L, Chiu W, De Jonghe S, Maes P, Slechten B, Raymenants J, et al. Remdesivir, Molnupiravir and Nirmatrelvir remain active against SARS-CoV-2 Omicron and other variants of concern. Antiviral Res 2022;198:105252.

[3] Painter GR, Natchus MG, Cohen O, Holman W, Painter WP. Developing a direct acting, orally available antiviral agent in a pandemic: the evolution of molnupiravir as a potential treatment for COVID-19. Curr Opin Virol 2021;50:17–22.

[4] Wahl A, Gralinski LE, Johnson CE, Yao W, Kovarova M, Dinnon KH 3rd, et al. SARS-CoV-2 infection is effectively treated and prevented by EIDD-2801. Nature 2021;591:451–7.

[5] Kumarasamy N, Jindal, A, Saha, B, Singh VB, Rodduturi NCR, Sinha S, Sriramadasu SC. Phase IIII trial of molnupiravir in adults with mild SARS-cov2 infection in India, 2022.

[6] Gordon CJ, Tchesnokov EP, Schinazi RF, Götte M. Molnupiravir promotes SARS-CoV-2 mutagenesis via the RNA template. J Biol Chem 2021;297:100770.

[7] Jayk Bernal A, Gomes da Silva MM, Musungaie DB, Kovalchuk E, Gonzalez A, Delos Reyes V, et al. Molnupiravir for oral treatment of Covid-19 in nonhospitalized patients. N Engl J Med 2022;386:509–20.

[8] Arribas JR, Bhagani S, Lobo SM, Khaertynova I, Mateu L, Fishchuk R, et al. Randomized trial of molnupiravir or placebo in patients hospitalized with covid-19. NEJM Evidence 2022;1. https://doi.org/10.1056/evidoa2100044.

[9] Flisiak R, Horban A, Jaroszewicz J, Kozielewicz D, Mastalerz-Migas A, Owczuk R, et al. Diagnosis and therapy of SARS-CoV-2 infection: recommendations of the Polish Association of Epidemiologists and Infectiologists as of November 12, 2021. Annex no. 1 to the Recommendations of April 26, 2021. Pol Arch Intern Med 2021;131. https://doi.org/10.20452/pamw.16140.

[10] Flisiak R, Horban A, Jaroszewicz J, Kozielewicz D, Mastalerz-Migas A, Owczuk R, et al. Management of SARS-CoV-2 infection: recommendations of the Polish Association of Epidemiologists and Infectiologists as of February 23, 2022. Pol Arch Intern Med 2022;132. https://doi.org/10.20452/pamw.16230.

[11] MI2DataLab. Monitor of SARS-CoV-2 n.d. https://monitor.crs19.pl/2022-03-31/poland/?lang=en. (accessed June 23, 2022).

[12] Flisiak R, Zarębska-Michaluk D, Berkan-Kawińska A, Tudrujek-Zdunek M, Rogalska M, Piekarska A, et al. Remdesivir-based therapy improved the recovery of patients with COVID-19 in the multicenter, real-world SARSTer study. Pol Arch Intern Med 2021;131:103–10.

[13] Zarębska-Michaluk D, Jaroszewicz J, Rogalska M, Martonik D, Pabjan P, Berkan-Kawińska A, et al. Effectiveness of tocilizumab with and without dexamethasone in patients with severe COVID-19: A retrospective study. J Inflamm Res 2021;14:3359–66.

[14] Wong CKH, Au ICH, Lau KTK, Lau EHY, Cowling BJ, Leung GM. Real-world effectiveness of molnupiravir and nirmatrelvir/ritonavir among COVID-19 inpatients during Hong Kong’s Omicron BA.2 wave: an observational study. BioRxiv 2022. https://doi.org/10.1101/2022.05.19.22275291.

